# The tissue-specific effects of glucose-lowering drug targets on aging mediated through DNA methylation: a multi-omics genetic study

**DOI:** 10.1101/2025.10.13.25337858

**Authors:** Yuqi Sun, Haonan Zheng, Min Ma, Rongrong Gu, Manqing Wang, Si Fang, Yangbo Sun, Qian Yang, Yufang Bi, Jie Zheng

**Author notes:** Correspondence to Yufang Bi, Qian Yang, and Jie Zheng. Equal contribution (Yuqi Sun, Haonan Zheng, Min Ma).

## Abstract

**Background:** DNA methylation plays a key role in mediating the anti-aging effects of glucose-lowering drugs. This study aims to systematically explore the potential anti-aging effects of target genes of FDA-approved glucose-lowering drugs and the underlying epigenetic mediators.

**Method:** We conducted a two-sample Mendelian randomization (MR) study to investigate the putative causal relationships between the gene expression levels of glucose-lowering drug targets and 10 aging-related phenotypes, followed by a two-step MR to estimate the mediation effect of DNA methylation. Drug candidates were selected according to the latest review of clinical drug use for type 2 diabetes, and their target genes were obtained from the DGIdb database. Tissue-specific *cis-*expression quantitative trait loci (eQTLs) from GTEx consortium were selected as genetic instruments to proxy the expression level of drug-target genes. Glycemic phenotypes were used as positive controls to validate the instruments. The *cis-* and *trans-*methylation QTLs of Cytosine-phosphate-Guanine sites near the drug target genes were obtained from GoDMC consortium. Additionally, we performed enrichment analyses focused on tissue specificity and aging pathways to further corroborate our findings.

**Results:** We obtained 194 target genes interacted with 36 FDA-approved anti-diabetic drugs, of which the tissue-specific eQTLs were used to proxy the drug target effects. MR showed strong evidence that 9 interacting genes of 6 glucose-lowering drugs showed anti-aging potential on one or more aging-related phenotypes mediated by DNA methylation: *EHMT2, HSPA4, IGF2BP2, IRS1, LPL, NDUFAF1, NDUFS3, SLC22A3* and *TCF7L2*. These genes were distributed in 17 tissues, especially in the central nervous system, suggesting a potential neural component in their anti-aging effects. For instance, expression of *EHMT2* in several brain basal ganglia regions, which the gene interacted with Tolazamide, showed a protective effect on frailty (odds ratio[OR] in caudate =1.02, 95%CI=1.01-1.04, FDR adjusted P=1.69×10^-2^; OR in putamen=1.02, 95%CI=1.01-1.03, P_FDR_=3.37×10^-2^, OR in nucleus accumbens=1.02, 95%CI=1.01-1.04, P_FDR_=3.37×10^-2^). These associations were externally validated by searching literature evidence in existing EWAS and TWAS studies, as well as evidence from enrichment analyses.

**Conclusions:** This study prioritizes nine glucose-lowering genes as anti-aging drug targets in specific tissues and prioritizes their epigenetic regulation through DNA methylation for future drug development.

## Introduction

Aging is a complicated process characterized by the progressive accumulation of molecular damage, where epigenetic alteration is considered one of the primary hallmarks and an unambiguous contributor [1]. Since the human DNA methylation (DNAm) landscape has been associated with plenty of aging-related phenotypes [2], specific therapeutic interventions on the epigenetic status yield tangible benefits for healthspan. Given the inconsistency of DNAm patterns across different human tissues, recent studies have emphasized the importance of developing tissue-specific interventions [3]. A retrospective review of randomized controlled trials (RCTs) summarized 8 categories of promising anti-aging compounds that act via various pathways, including epigenetic alteration [4]. Two of them are glucose-lowering drugs: metformin and glucagon-like peptide-1 receptor agonists, thus attracting people’s attention to the anti-aging potential of all types of glucose-lowering drugs.

RCTs involving glucose-lowering drugs are usually designed to evaluate their effects on age-associated diseases rather than aging [5]. Those RCTs were designed to recruit high-risk participants (e.g., those with cardiovascular diseases or dementia), thus providing limited evidence of extending healthspan or decelerating aging in the general population [5]. A small RCT, Metformin in Longevity Study (N = 14), provided preliminary evidence that metformin can lead to transcriptomic changes in pathways affecting aging in skeletal muscle and subcutaneous adipose tissues [6]. An ongoing RCT, Targeting Aging with Metformin [7], aims to explore the effect of metformin among 3,000 ethnically diverse, non-diabetic participants aged 65–80, but the results have not been released yet. Besides, only a few clinical trials explored the beneficial effects of other glucose-lowering agents on aging, including acarbose (N = 10, NCT02953093), canagliflozin (N = 30, NCT06301529), and dapagliflozin (N = 20, NCT04401904), all of which have very limited sample sizes.

Given the limitations of conventional observational studies and inconclusive results from RCTs, human genetics could help shed new light on the anti-aging nature of glucose-lowering drug targets, which may further facilitate the development of new lifespan-extending medications [8]. Previous Mendelian randomization (MR) studies indicated potentially protective effects of targets for metformin, sodium-glucose cotransporter 2 inhibitors, and sulfonylureas on aging-related diseases, including cardiovascular diseases, cancer, dementia, inflammation, and heart failure [9-12]. In addition, metformin might promote healthy aging via targeting GPD1 and AMPKγ2 (*PRKAG2*) [10]. Nevertheless, it remains unclear whether glucose-lowering drugs protect against aging itself rather than aging-related diseases, and whether they function through epigenetic alterations.

The aim of this study is to explore the target-specific effects of glucose-lowering drug targets on aging biomarkers using two-sample MR. To provide mechanistic insights, we further used two-step MR to investigate whether the effect of glucose-lowering drug targets on aging is mediated through DNAm.

## Methods

### Study design

Fig.1 presents the study design and workflow, including a schematic representation detailing the research procedure adopted and a graphical abstract outlining the data resource and core assumptions of the study.

**Figure 1.**
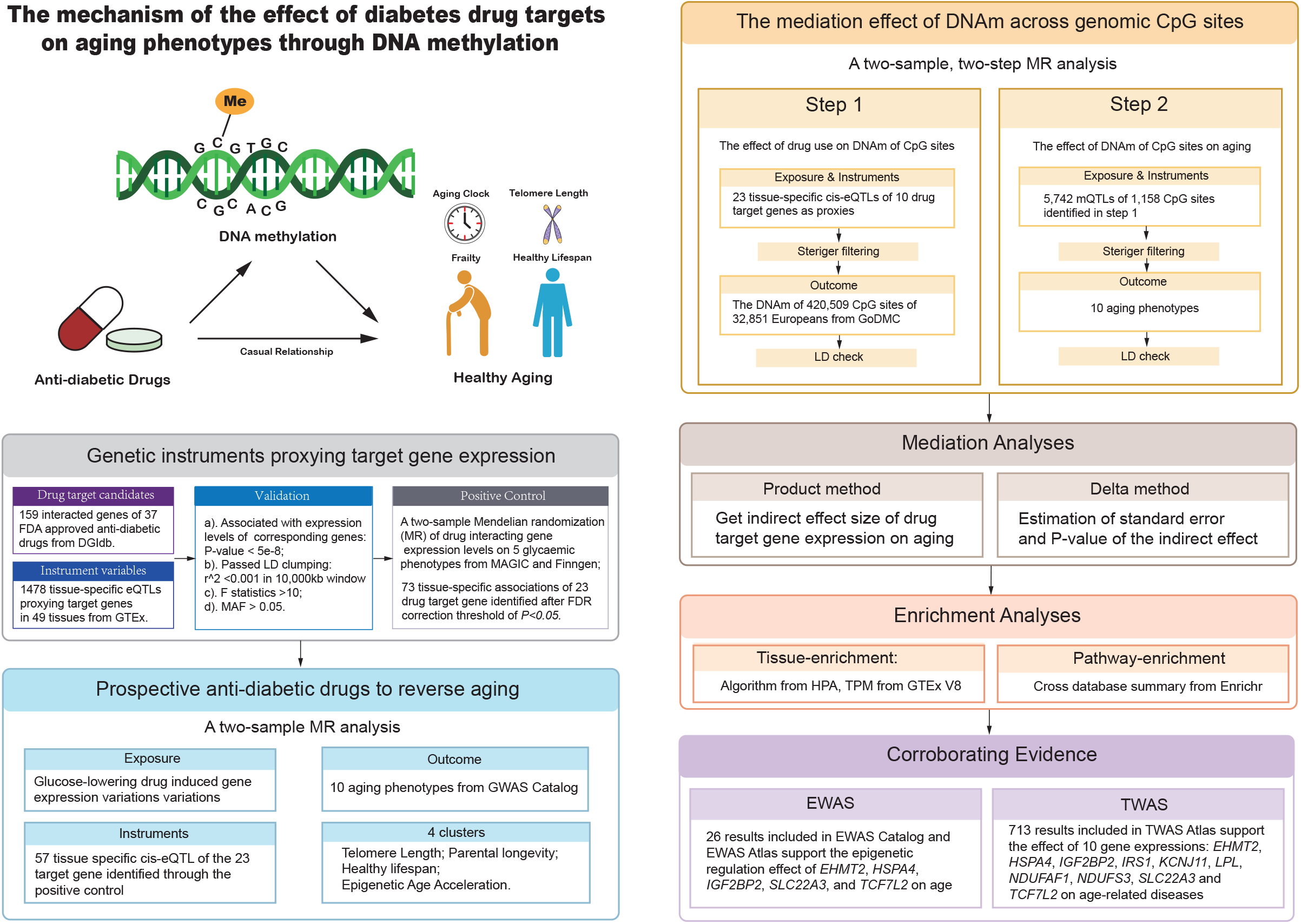
The study design and workflow. A schematic representation detailing the research procedure adopted and a graphical abstract outlining the core assumptions of this study.

### Selection of genetic instruments for glucose-lowering drug targets

This study considered nine major categories of glucose-lowering drugs used in first-line clinical practice [13]: Glucagon-Like peptide-1 receptor agonists, biguanides, sulfonylureas, meglitinides, α-glucosidase inhibitors, dipeptidyl peptidase-4 inhibitors, sodium-glucose cotransporter 2 inhibitors, thiazolidinediones, and Lck/Yes-related Novel protein tyrosine kinase activators. A total of 36 FDA-approved drugs were selected as candidates. Since the mechanism by which these drugs act is not fully understood, we searched the listed glucose-lowering drugs in the Drug-Gene Interaction database (DGIdb) v.5.0.7 [14], and reserved 194 related genes as potential targets. Using information implemented in DrugBank, those gene targets with clear mechanisms of action were prioritized as Tier 1 druggable genes, where those genes exhibiting drug-gene interactions were classified as Tier 2 (electronic supplementary material [ESM] Table 1).

We obtained tissue-specific *cis-*acting expression quantitative trait loci (*cis-*eQTLs) of each glucose-lowering drug target gene from Genotype-Tissue Expression (GTEx) v8 Project (15,201 samples of 49 tissues from 838 donors), and used them to select tissue-specific genetic instruments [15]. A stringent selection process was performed to identify tissue-specific genetic variants for each target, requiring a SNP a) being associated with expression levels of corresponding genes (*P* < 5 × 10^-8^) in the *cis*-acting regions (i.e. ±500 kb window of each gene probe); b) passing a stringent linkage disequilibrium (LD) clumping test using the panel of European ancestry with clumping *r*^2^ of less than 0.001 and clumping window of 10,000 kb; c) having an F-statistic > 10, which indicating good instrument strength; d) having minor allele frequency ⍰≥ 0.01. We also applied Steiger filtering to identify the genetic variants explaining more variations of the exposures than the outcomes, thereby addressing potential reverse causality [16]. In addition, we considered five glycemic traits: Hemoglobin A1c, fasting glucose, fasting insulin, 2-hour glucose, and type 2 diabetes (T2D) as positive controls to validate the reliability of the instruments. The associations of genetic susceptibility to the targets with glycemic traits were estimated using two-sample MR. Only targets relevant to at least one of these traits (false discovery rate, FDR < 0.05) were reserved for further analyses.

### Selection of aging proxy indicators

A variety of aging biomarkers and phenotypes have been proposed for the identification and evaluation of longevity interventions [17]. Referring to the latest aging biomarker consensus and previous MR studies about aging [17, 18], we selected ten phenotypes as aging proxy indicators: father’s age at death, mother’s age at death, combined parental age at death, Hannum Age, Intrinsic Horvath Age, PhenoAge, Grimage, leukocyte telomere length, healthspan, and frailty. The details of genome-wide association studies (GWAS) on each phenotype are described in ESM Method 1. The main characteristics of each GWAS are presented in Supplementary Table 1.

### Selection of DNAm mediators

The GoDMC study published a database of > 270,000 independent DNAm quantitative trait loci (mQTLs), involving 32,851 European participants [19]. Putative genetic variants associated with DNAm at 420,509 CpG sites were identified from blood samples, with genotype data imputed to the 1000 Genomes Project reference panel [20], and DNAm profiles quantified from bisulfite-converted genomic whole-blood DNA using the Infinium HumanMethylation BeadChip (HumanMethylation450 or EPIC arrays). We localized the genome-wide DNAm datasets and the genetic location of all CpG sites available from GoDMC to establish a retrieval library, using a standardized pipeline to convert all data to hg19/GRCh37 build to ensure consistency. For genetic instruments of CpG sites, we used the same standard applied in eQTL instrument selection. CpG sites associated with expression levels of the drug-related genes as identified by MR results, and located within ± 500kb of the gene, are considered *cis*-regulated by drug-related targets; otherwise, they are classified as *trans*-regulated.

### Selection of positive controls

To ensure our findings reflect specific pharmacological effects rather than non-specific processes such as drug metabolism, we restricted our analyses to target genes that exhibited a significant impact on glycemic traits. GWAS of five indexes which reflect the change of blood glucose are used as outcomes in positive controls to select valid instruments for drug targets, including HbA1c level (N = 146,806), fasting glucose level (N = 200,622), fasting insulin level (N = 151,013), 2-hour glucose level (N = 63,396) from MAGIC consortium [21] and T2D (N = 1,812,017, N cases = 242,283, N controls = 1,569,734) from the T2DGGI study [22]. Participants were of European descent, and detailed information on these GWAS is summarized in Supplementary Table 2.

### Statistical analysis

#### Genetic correlation analysis

We used LD-score regression to calculate the genetic correlations and SNP heritability of the 10 aging traits [23-25], based on the European-ancestry LD reference panel. In line with recommendations [25], low-quality (INFO⍰< ⍰0.9) and rare (minor allele frequency⍰< ⍰0.01) SNPs, as well as those located in the Major Histocompatibility Complex, were discarded before merging the summary statistics. We further clustered those ten traits into four groups according to their genetic correlations.

#### Mendelian randomization analyses

We firstly conducted a two-sample MR to identify the glucose lowering drug target genes whose expression level causally affect the aging outcomes. A two-step MR was further conducted to assess whether DNAm could mediate the effect. The first step is to assess how each target gene’s expression level influences DNAm of CpG sites across the whole genome. The second step was carried out on those DNAm that passed the first step (MR P_FDR_< 0.05). For these DNAm, we evaluated whether they affect aging-related outcomes. Mediation analysis was then performed to identify the mediation proportion of the DNAm of CpG sites in each target gene expression and aging outcome pair. The product of coefficients method and the delta method were applied to calculate the indirect effect and the standard error of the indirect effect, respectively, for each of the mediator [26].

For all the MR analysis listed above, we utilized Wald’s ratio method for exposures with only one instrument, and the inverse variance weighted (IVW) method for exposures with two or more instruments [27]. To ensure the robustness of associations, we further performed sensitivity analyses using weighted median and weighted mode methods for exposures with three or more instruments. For instruments that are not available in the outcome GWAS data sets during the MR analysis, proxy genetic variants in high LD with this eQTL/mQTL (*r*^*2*^ > 0.8 in the 1000 Genomes data in the European population [20]) were used as substitutes.

#### Validation of MR assumptions

MR analyses require three key assumptions: (1) relevance: the genetic predictors have robust associations with the exposure; (2) exchangeability: there are no common causes (confounders) between the instrument and outcome; (3) exclusion restriction: the instruments are only related to the outcome through the exposure being studied [28]. Findings were reported according to the STROBE-MR (Strengthening the Reporting of Mendelian Randomization Studies) guidelines (ESM Method 2) [29].

To assess the relevance assumption, we examined the strength of each genetic variant using F-statistics. Only variants meet that this standard will be considered valid instrumental variables.

The exchangeability assumption cannot be definitively proven, but associations with known confounders can be tested and demonstrated [30]. We used GWAS in European participants to avoid population mixture. These GWAS were adjusted for the top principal components or through the use of adjusted linear mixed models to minimize the confounding by population stratification [31]. We also conducted LD check analysis to evaluate the LD structures of causal genes or CpGs with robust MR evidence (FDR < 0.05) and mitigate bias resulting from confounding by LD [32]. We estimated the pairwise LD between each eQTL/mQTL and mQTL/aging-associated GWAS variant in the *cis-*region. We utilized the threshold of LD *r*^*2*^ > 0.8 as evidence of LD checked for the whole study [32]. Only results that pass the LD check will be included in further analysis.

To assess the exclusion restriction assumption, we applied two sensitivity tests: weighted median and weighted mode methods, to minimize potential bias and improve confidence in our causal interpretation when more than 3 instruments were available.

#### Enrichment analyses

We performed tissue-enrichment analyses for all prospective drug target genes identified through the MR analyses. Gene-level RNA-Seq expression values, measured in transcripts per million (TPM), were obtained from the GTEx V8 release. The tissue-specific genes are defined using the simplified algorithm from the Human Protein Atlas (ESM Method 2) [33]. We also queried Enrichr to identify the tissue-specific aging-associated expression variation and relevant aging pathways for the drug target genes [34-36].

#### Triangulation of evidence

Public databases of epigenome-wide association study (EWAS) and transcriptome-wide association study (TWAS) were reviewed to support our discoveries. We searched the EWAS Catalog (retrieval version: data last updated on March 22, 2024) [37] on the EWAS Open platform, to explore DNAm associations for our drug target genes. We also searched the TWAS Atlas (retrieval version: data last updated on May 15, 2022) [38], to identify disease associations with gene transcription in whole blood and multiple tissues, collecting tissue-specific TWAS results to further support our findings.

## Results

### Summary of instrument and outcome selection

We searched drug-gene interaction (DGI) for 36 glucose-lowering drugs (Fig. 2, ESM Table 1). A total of 194 drug-related genes were selected as candidates; 160 of them were found in the DGI database with 1,619 corresponding *cis*-eQTLs from 49 types of human tissues identified as instruments (ESM Table 2). Among these, the expression of 23 genes showed a causal effect on at least one glycemic phenotype in tissue-specific conditions at the FDR threshold of P < 0.05 and were regarded as drug target genes for this study (Supplementary Fig. 1, ESM Table 3).

**Figure 2.**
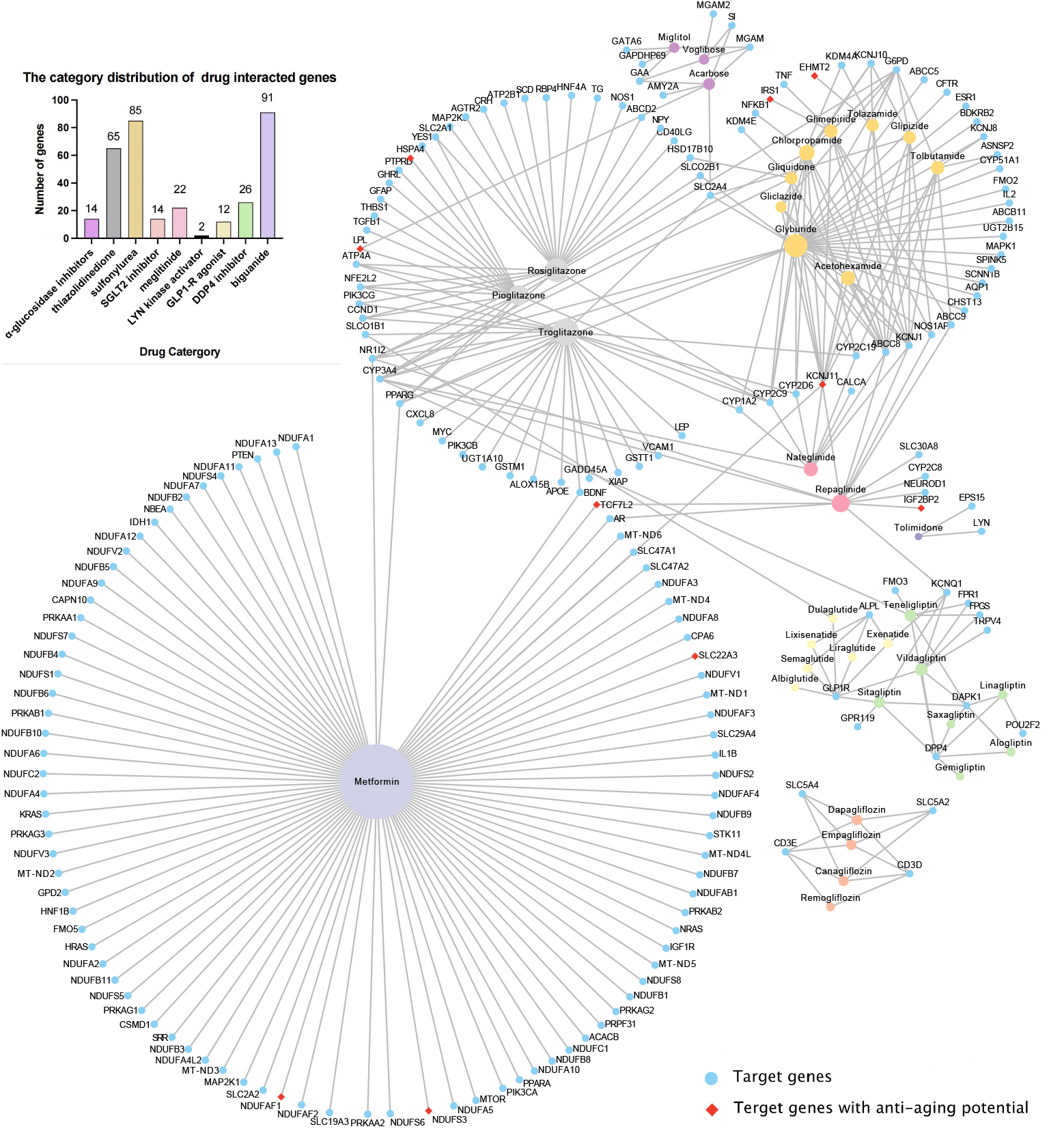
Category map of first-line glucose-lowering drugs and their interacted genes. Genes that showed a causal relationship with aging phenotypes are marked with red diamonds. The bar chart in the upper left corner shows the distribution of target gene categories. In total, this study collected 36 FDA-approved glucose-lowering drugs across 9 categories: 14 α-glucosidase inhibitors, 65 thiazolidinediones, 85 sulfonylureas, 14 SGLT-2 inhibitors, 22 meglitinides, 2 LYN kinase agonists, GLP-1 receptor agonists, DPP-4 inhibitors, and 91 biguanides.

By calculating genetic correlations (rg) and subsequently performing hierarchical clustering based on the magnitude of these correlations, we found that the 10 aging phenotypes from four distinctive clusters of high genetic similarity (|rg|⍰≥ ⍰0.2; P⍰< ⍰5×⍰10^-3^) (Supplementary Fig. 2, ESM Table 4). Thus, we categorized them into four groups: telomere length, parental longevity, healthspan and epigenetic age acceleration (EAA), to represent aging from different aspects.

### Tissue-specific associations of gene expression levels of glucose-lowering drug targets with aging outcomes

Through the two-sample MR, ten target genes (*EHMT2, HSPA4, IGF2BP2, IRS1, LPL, NDUFAF1, NDUFS3, SLC22A3, TCF7L2*, and *KCNJ11*) of sixteen drugs (acarbose, acetohexamide, canagliflozin, chlorpropamide, dapagliflozin, gliclazide, glimepiride, glipizide, gliquidone, glyburide, metformin, nateglinide, pioglitazone, repaglinide, tolazamide and tolbutamide) displayed strong evidence for their tissue-specific expressions to be causally related to aging phenotypes (Fig. 3, ESM Table 5). We found the expression levels of *EHMT2, HSPA4*, and *NDUFS3* in multiple brain regions, including caudate basal ganglia, cerebellar hemisphere, nucleus accumbens basal ganglia, and putamen basal ganglia, were associated with frailty and leukocyte telomere length. *LPL* and *SLC22A3* expression in the tibial nerve was associated with parental age (odds ratio [OR]=1.06, 95%CI=1.03-1.09, P_FDR_=3.36×10^-3^) and health span (OR=1.07, 95%CI=1.02-1.11, P _FDR_=3.37×10^-2^).

**Figure 3.**
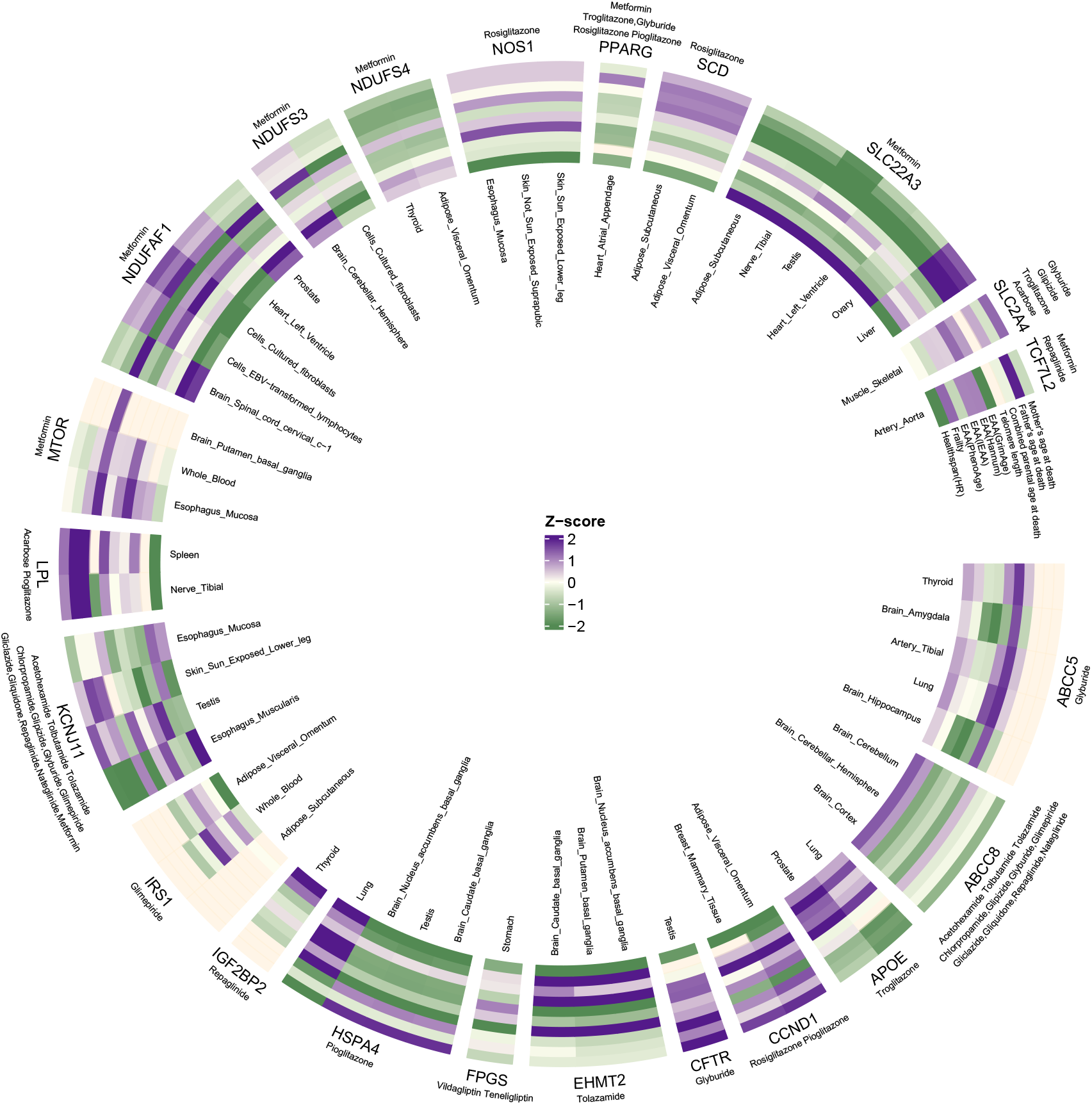
Circular plot showing all MR estimations of drug target genes’ expression levels on aging phenotypes. Z-scores were calculated as the effect size divided by its standard error (β/SE) to represent the strength and direction of each association. The innermost ring indicates the tissue of origin for each gene’s expression, while the outermost ring shows the name of the drug associated with each gene. EAA, epigenetic age acceleration. HR, hazard ratio. Detailed MR results are provided in ESM Table 5.

In addition, *SLC22A3* also exhibited a pan-tissue effect, which showed associations with healthspan in multiple tissues, including the left ventricle (OR=1.05, 95%CI=1.02-1.08, P _FDR_=3.22×10^-2^), liver (OR=0.96, 95%CI=0.95-0.98, P_FDR_=6.29×10^-4^), subcutaneous adipose tissue (OR=1.06, 95%CI=1.02-1.10, P_FDR_=6.29×10^-4^), ovary (OR=1.03, 95%CI=1.02-1.05, P_FDR_=1.03×10^-2^), and testis (OR=1.06, 95%CI=1.04-1.09, P _FDR_=1.42×10^-4^) in addition to tibial nerve.

### Mediation analysis

In Step 1 of the two-step MR, associations of 3,526 pairs of target genes and CpG sites were tested, with 3,230 showing at least marginal MR evidence, including 2,064 *cis-*acting associations and 1,166 *trans-*acting associations (ESM Table 6, Fig. 4). In Step 2, the 5,742 genetic variants robustly associated with DNAm were used as instruments to proxy 1,158 CpG sites. We then tested the associations between DNAm of CpG sites and aging traits (ESM Table 7 and 8, Supplementary Fig. 3). Eventually, nine target genes (*EHMT2, HSPA4, IGF2BP2, IRS1, LPL, NDUFAF1, NDUFS3, SLC22A3* and *TCF7L2*) of eight drugs: acarbose, canagliflozin, dapagliflozin, glimepiride, metformin, pioglitazone, repaglinide and tolazamide, were associated with aging outcomes mediated by DNAm of 1,058 CpG sites (ESM Table 9, Fig.5, Supplementary Fig. 4). 755 of 1,058 (71.4%) of the mediating DNAm are *cis-*regulated by their corresponding drug target genes, except *EHMT2* exhibits both *cis-* and *trans-*regulated DNAm. The target genes mediated through DNAm distribute in 16 tissues of five systems, including nervous system (caudate, nucleus accumbens, putamen and cerebellar hemisphere and tibial nerve), cardiovascular system (artery aorta and left ventricle), immune and endocrine system (cultured fibroblasts, EBV-transformed lymphocytes, spleen, adipose tissues, liver and thyroid), respiratory system (lung) and reproductive system (testis and ovary).

**Figure 4.**
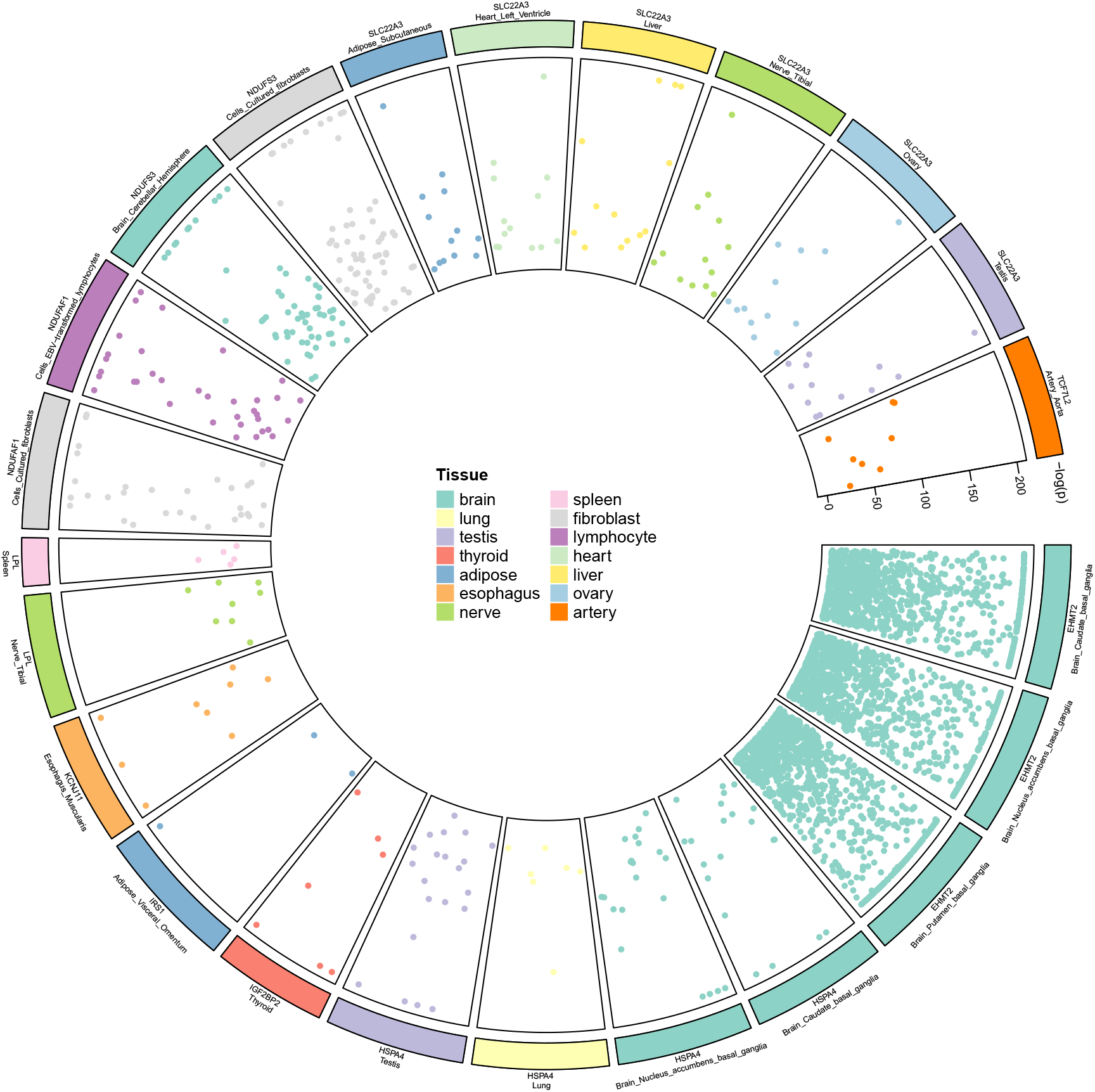
Circular plot showing CpG sites distribution associated with the target genes in relevant tissues. Each spot represents a single CpG site. The y-axis shows the −log_10_-transformed P_FDR_-values indicating the statistical significance of the association between genes and CpG site methylation levels in each tissue. Detailed MR results are provided in ESM Table 6.

**Figure 5.**
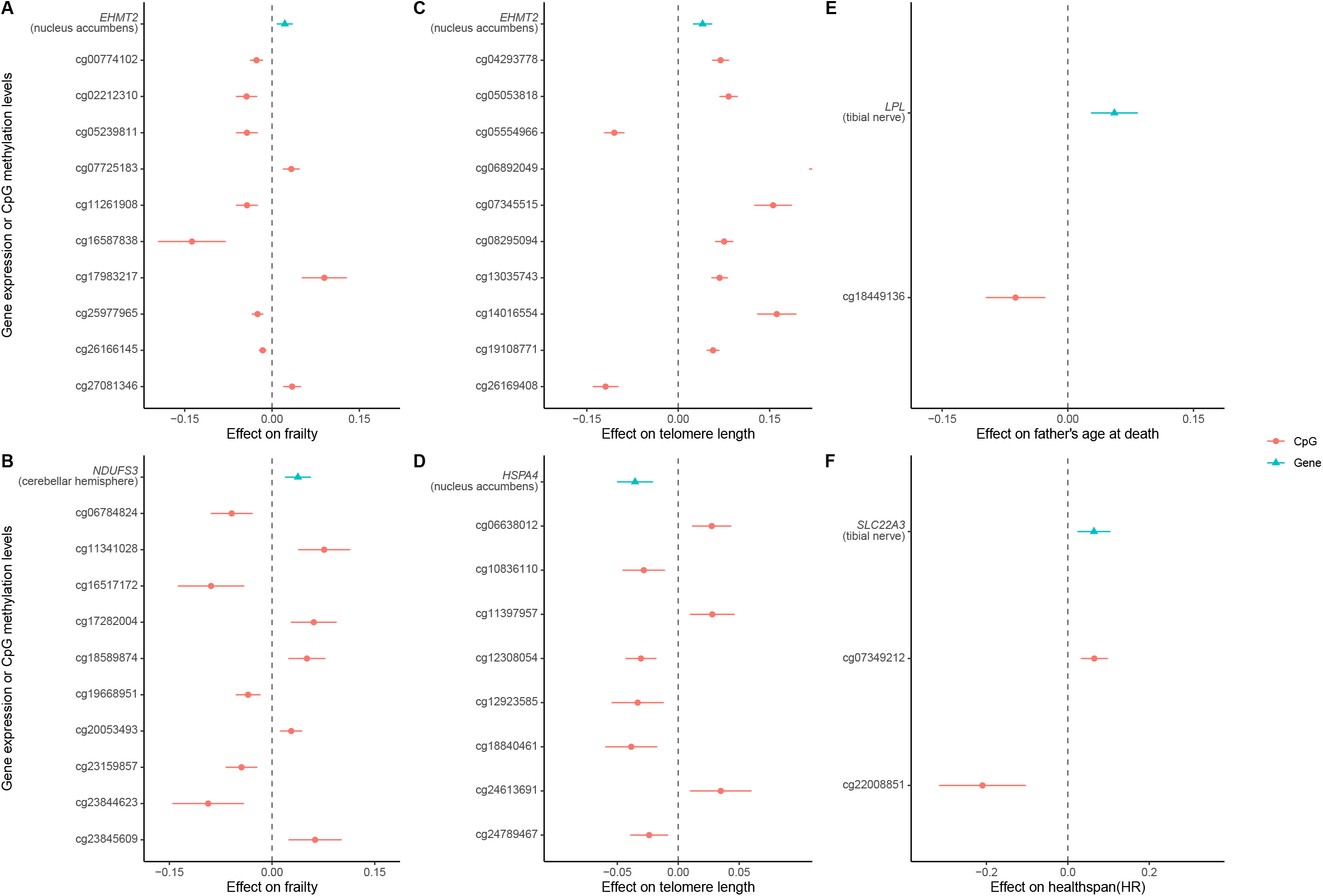
Forest plot showing MR results of tissue-specific expression levels of drug target genes on aging phenotypes, along with their associated mediating CpG sites. 5 genes were selected as representative examples based on prior knowledge and are indicated at the top of each panel. The top half of each chart displays the effect of gene expression on aging outcomes. The x-axis represents the change in outcome per standard deviation (SD) increase in the expression level of each drug target gene. The bottom half of each chart shows the top cis-CpG site mediators (up to 10 per gene), selected based on the lowest p-values of the mediation effect. The x-axis represents the change in the outcome per standard deviation (SD) increase in the methylation level of each CpG site. Detailed estimations are provided in ESM Table 5, 7, 8 and 9.

### Triangulation of external evidence

Results extracted from the previous EWAS study supported the mediation effect of relevant target genes and aging via epigenetic regulation. DNAm of five genes: *EHMT2, HSPA4, IGF2BP2, SLC22A3*, and *TCF7L2*, showed associations with aging-related outcomes in EWAS, which in line with our findings (ESM Table 10). For TWAS findings reported in TWAS Atlas, only a limited number of studies reported associations with aging-related outcomes. Thus, the top eight aging-related diseases, as well as their common risk factors, were included to proxy aging, in accordance with the definition of healthspan phenotype [39]. All ten prioritized aging associated genes also showed relevant TWAS evidence: *EHMT2, HSPA4, IGF2BP2, IRS1, LPL, NDUFAF1, NDUFS3, SLC22A3* and *TCF7L2* (ESM Table 11). Tissue enrichment analysis showed no evidence of preferential expression of drug target genes in tissues corresponding to MR results (Supplementary Table 3). However, results from Enrichment analysis from Enrichr indicated that these genes are members of several metabolism, mitochondrial functions, insulin/IGF-1 signaling pathways and other aging-associated pathways [40, 41], with tissue-specific regulatory relevance of aging further supported by external evidence from GEO datasets [41].

## Discussion

This drug target genetic study comprehensively investigated the tissue-specific effect of glucose-lowering drug targets on a range of aging phenotypes mediating through DNAm. We extended the recent discoveries of anti-aging targets and identified DNAm at multiple CpG sites as potential mediators in the aging process, highlighting the importance of exploring epigenetic regulatory mechanisms in aging-related therapies. Our study showed potentials of eight relevant anti-diabetic drugs as prospective candidates to resist aging. The tissue-specific expression of nine drug-targeting or drug-interacting genes: *EHMT2, HSPA4, IGF2BP2, IRS1, LPL, NDUFAF1, NDUFS3, SLC22A3*, and *TCF7L2* as targets for future research on decelerating aging with a focus on their epigenetic regulations through DNAm.

Among the eight anti-diabetic drugs, acarbose [42], canagliflozin [43], dapagliflozin [44] and metformin [45, 46], and pioglitazone [47] have previously shown the potentials to extend lifespan or decrease the risk of several aging-related diseases in animal models. However, their evidence in humans was largely based on observational studies [6, 48-51], leaving it unclear whether these associations are driven by residual confounding, reverse causality and immortal time bias [52]. Besides, most of these studies focused on the protective effects of the anti-diabetic drugs in high-risk populations from certain aging-related diseases (e.g. heart failure) rather than aging directly. Our results provided evidence to support the conservatism of the anti-aging effects of the relevant drug targets in the general population. Among the drugs we identified, glimepiride, repaglinide and tolazamide showed their potential of extending lifespan that were not reported before. These drugs could be considered as drug candidates for future clinical research.

The tissue-specific effects of gene expression we observed align well with previous studies of age-related diseases. For instance, low-dose pioglitazone has been shown to extend lifespan and improve cognitive function in aged mice by modulating metabolic homeostasis [53-55], though no direct evidence has been reported in humans. In this study, we identified the mitochondrial protein *HSPA4*, as a potential core gene in pioglitazone’s regulatory pathway of decelerating aging in brain tissues. Notably, *HSPA4* expression has also been found to be downregulated in human front cortex as well as in several mouse brain tissues [41], consistent with the tissue specificity highlighted by our MR results. In addition, a previous study found that acarbose has a pronounced lifespan-extending effect in male mice [42]. Here, we found one of its target genes, *LPL*, and its expression in the tibial nerves is the only exposure related to parental longevity, particularly the father’s life span. Similarly, *SLC22A3* expression in testis has been found to be related to combined parental age at death, which aligned with the gender difference in metformin effect on aging [46]. This may contribute as a starting point to further investigate the gender difference in the anti-aging effect of drugs.

We uncovered potential anti-aging roles of target genes for two sulfonylurea drugs (*EHMT2* for tolazamide and *IRS1* for glimepiride), and a meglitinide class drug (*IGF2BP2* for repaglinide). Specifically, *EHMT2* is a well-recognized histone methyltransferase to have the ability of regulating DNAm [56]. It is also a member of the Senescence Associated Secretory Phenotype (SASP) pathway [56], and has been observed to be downregulated in aging mouse hippocampus. Results from model organisms suggest *IRS1* is involved in the IGF1/GH1 axis that could regulate aging [57], with loss of expression extending lifespan in mice [58] and *Drosophila* [59]. *IGF2BP2* has been identified as a key factor to activate stem cell aging [60] and brain aging [61] in mouse models, and also recognized as an epigenetic regulatory factor in epitranscriptomics [62]. This warrants further investigation to clarify its roles in human aging, including its involvement in the insulin/IGF signaling pathways and the crosstalk between RNA and DNA modification. Metformin has been shown in multiple studies to exert lifespan-extending effects [48]. Here we identified four novel target genes: *NDUFS3, NDUFAF1, SLC22A3*, and *TCF7L2*, mainly involved in mitochondrial and energy metabolism pathways [40], that may underlie Metformin’s novel anti-aging mechanisms and merit further investigation. Notably, three of them: *EHMT2, HSPA4* and *NDUFS3*, showed a brain-specific effect on aging, which were mediated by methylation variation across multiple CpG sites. This underscores the key role of epigenetic changes in the connecting central nervous system regulation and systemic aging.

The strengths of this study included the widely used pipelines for instrument selection and a high resolution of tissue-specific results. In addition, we included a comprehensive set of aging outcomes, a wide range of drug target selection, and an emphasis on epigenetic regulation of aging. We also integrated external multi-omics evidence to enhance the interpretations of our findings, aiming to provide more informative and predictive results compared with previous drug-target MR studies on aging and offering implications for future targeted drug development [11, 18, 63, 64].

This study still has limitations. First, the eQTL data used in this study are derived from a relatively small sample size of 838 individuals, with only one instrument available for each target gene per tissue. In addition, solely relying on lead cis-eQTLs for target genes in each tissue may limit the detection of potential bias [30, 65]. Second, some target genes, such as *TCF7L2*, have qualified instruments in only a few tissues. This led to the incomplete exploration of the tissue-specific function of these genes. Third, the data for DNAm, the EAA group of aging indicators, and leukocyte telomere length were derived from blood samples, introducing bias due to sample incoherence. Fourth, for drugs like acarbose [42] and canagliflozin [43], the utilization of sex-stratified data, including sex-stratified eQTL data and sex-stratified GWASs of aging phenotype, may help elucidate gender-specific differences in epigenetic regulations [66]. Fifth, not all aging hallmarks listed in the consensus have publicly available GWAS datasets [1]. Selection bias is inevitable in our study, which may contribute to the results that no gene was found to affect the epigenetic age acceleration group. More GWAS meta-analyses are required for better investigation. Sixth, several CpG sites showed a mediation proportion exceeding 100%. One of the three following conditions accounts for the phenomenon: (1) there are other mediators with a negative portion mediated; (2) the DNAm of each CpG site affects one another; (3) there are interactions between the DNAm of each CpG site [67, 68]. Thus, the observed indirect effect requires further investigation. Seventh, the intervention of these drugs could have varying effects depending on age groups, ethnic backgrounds, and disease states. We need to assess the specific effects of these drug-gene expressions across diverse populations to understand the generalizability of our results in the near future. Finally, MR can only proxy the drug target effect rather than the direct effect of the drug on aging. Therefore, the evidence provided by this study cannot be used for any clinical investigations before formal clinical trials.

## Conclusion

In conclusion, our study identified 9 target genes of 6 glucose-lowering drugs showing robust effects on longevity through DNA methylation. We identified both previously recognized and novel candidate genes that may contribute to lifespan extension by modulating aging-associated pathways, such as SASP signaling, mitochondrial and lipid metabolism, insulin signaling, and epitranscriptomic regulation. Several effects were tissue-specific, particularly within Five drug interacting genes exhibited a tissue-specific enrichment in the nervous system, emphasizing the neural regulation in systemic aging. Previous studies provide strong support for these findings. Further clinical and experimental research is required to validate and explore the feasibility and effectiveness of targeting these genes and drugs for aging prevention.

## Data Availability

All data produced in the present study are available upon reasonable request to the authors

## Abbreviations

DNAm: DNA methylation
EAA: Epigenetic age acceleration
EWAS: Epigenome-wide association study
FDR: False discovery rate
eQTL: Expression quantitative trait locus
GWAS: Genome-Wide Association Study
LD: Linkage disequilibrium
mQTL: DNAm quantitative trait locus
MR: Mendelian randomization
RCT: Randomized controlled trial
TWAS: Transcriptome-wide association study
T2D: Type 2 diabetes

## Declarations

### Availability of data and materials

The GWAS summary statistics for the ten primary aging outcomes are available from the GWAS Catalog database (www.ebi.ac.uk/gwas/). The GWAS summary statistics for glycemic traits are contributed by MAGIC investigators (www.magicinvestigators.org) and T2DGGI study (http://www.diagram-consortium.org/downloads.html). The analytical script of the MR analyses conducted in this study is available via the GitHub repository of the ‘TwoSampleMR’ R package (github.com/MRCIEU/TwoSampleMR/). The analytical script of the genetic correlation analyses conducted in this study is available via the GitHub repository of the ‘ldsc’ R package (github.com/mglev1n/ldscr/). The code used during the current study is available upon reasonable request from the corresponding author. All the other data generated or analysed during this study are included in this published article and the ESM files.

### Competing Interest

J.Z and Y.B.S is the editorial board member of BMC medicine. Other authors declare that they have no competing interests.

### Funding

This study was supported by grants from the Noncommunicable Chronic Diseases-National Science and Technology Major Project (2024ZD0531500, 2024ZD0531502), Students’ Innovation Training Program, Shanghai Jiao Tong University, School of Medicine, China (1824955Y) and grants from the National Key Research and Development Program of China (2022YFC2505203). Y.Q.S is supported by Roche University Research Fund, Chinese Society of Biochemistry and Molecular Biology, China (20240513). Z.J. and Q.Y. are supported by the National Natural Science Foundation of China (32570728 and 32500519). Y.B. is supported by the Shanghai Municipal Education Commission–Gaofeng Clinical Medicine Grant Support (20161307 and 20152508 Round 2).

### Contribution Statements

Y.Q.S., H.Z. and J.Z. conceived and designed the study. Y.Q.S drafted the manuscript. H.Z performed data analysis. M.M., S.F., Y.B.S., Q.Y. and J.Z. contributed to reviewing and editing the manuscript. Y.Q.S. and H.Z. contributed to visualization. Y.Q.S., H.Z., R.G. and M.W. contributed to data management and related paperwork. Y.Q.S. and H.Z. accessed and verified the data reported in the study. J.Z. is the guarantor of the findings. All the authors approved the final version to be published.

## Acknowledgments

We thank all of the investigators and participants who provided these data to support this study. We thank Huiling Zhao, Haoyu Liu and Xueyan Wu for the help of device acquisition and instructions.

## Author’s relationship and activities

Y.Q.S., H.Z., R.G. and M.W. are members of the College Innovation Training Program in Shanghai Jiao Tong University, School of Medicine. All authors declare that there are no relationships or activities that might bias, or be perceived to bias this work.

## Legends for figures

**Supplementary Figure 1** Forest plot of the top MR results: the effects of drug target gene expression levels on glycemic phenotypes. Outcomes include 2-hour glucose (A), HbA1c (B), fasting glucose (C), fasting insulin (D), and type 2 diabetes (E). The x-axis represents the change in the outcome per standard deviation (SD) increase in the expression level of each drug target gene. Statistical significance was defined as P_FDR_< 0.05. Detailed MR estimates are provided in ESM Table 3.

**Supplementary Figure 2** Genetic correlations between ten aging outcomes. Diagonal values show the observed-scale SNP heritability of each aging-related phenotype and off-diagonal circles show the genetic correlation among these traits. Blank squares are those did not pass multiple testing correction threshold (FDR⍰> ⍰0.05). The bottom dendrogram shows the hierarchical relationship between traits, based on the magnitude of their genetic correlations. The black box highlights the 4 clusters of aging-related phenotypes used in follow-up analyses: telomere length, parental longevity, healthspan and epigenetic age acceleration (EAA). HR, hazard ratio. Detailed results are provided in ESM Table 4.

**Supplementary Figure 3** Genomic distribution of mQTL and the target genes. The bar chart (A) on the top represents the position of target gene candidates and the count of CpG sites associated with them. The scatter plot (B) below represents the genomic positions of each mQTL and its corresponding CpG site. Detailed results are provided in ESM Table 6.

**Supplementary Figure 4** Forest plot of all MR results of the expression level of drug target genes on ten aging phenotypes, including combined parental age at death (A), father’s age at death (B), frailty (C), healthspan (D), leukocyte telomere length (E). The horizontal axis represents the effect size of exposure on outcomes. HR, hazard ratio. Statistical significance was defined as P_FDR_< 0.05. Detailed MR estimations are provided in ESM Table 5.

## Notes

### Competing Interest Statement

The authors have declared no competing interest.

### Author Declarations

The study used ONLY openly available human data that were originally located at: GTEx (https://www.gtexportal.org/); GoDMC (http://www.godmc.org.uk/);GWAS Catalog database (www.ebi.ac.uk/gwas/);MAGIC investigators (www.magicinvestigators.org); T2DGGI study (http://www.diagram-consortium.org/downloads.html)

